# Reactogenicity Differences between COVID-19 Vaccines: A Prospective Observational Study in the United States and Canada

**DOI:** 10.1101/2024.06.25.24309259

**Authors:** Matthew D. Rousculp, Kelly Hollis, Ryan Ziemiecki, Dawn Odom, Anthony M. Marchese, Mitra Montazeri, Shardul Odak, Laurin Jackson, Hadi Beyhaghi, Seth Toback

**Affiliations:** Novavax, Inc. Gaithersburg, MD 20878, USA; RTI Health Solutions, Research Triangle Park, NC 27709, USA

**Keywords:** Booster, COVID-19, reactogenicity, real-world evidence, SARS-CoV-2, NVX-CoV2373, mRNA

## Abstract

Participants in studies investigating COVID-19 vaccines commonly report reactogenicity events, and concerns about side effects may lead to reluctance to receive updated COVID-19 vaccinations. A real-world, post hoc analysis, observational 2019nCoV-406 study, was conducted to examine reactogenicity within the first 2 days after vaccination with either a protein-based vaccine (NVX-CoV2373) or an mRNA vaccine (BNT162b2 or mRNA-1273) in individuals who previously completed a primary series. Propensity score adjustments were conducted to address potential confounding. The analysis included 1130 participants who received a post–primary series dose of NVX-CoV2373 (n = 303) or an mRNA vaccine (n = 827) during the study period. Within the first 2 days after vaccination, solicited systemic reactogenicity events (adjusted) were reported in 60.5% of participants who received NVX-CoV2373 compared with 84.3% of participants who received an mRNA vaccine; moreover, 33.9% and 61.4%, respectively, reported ≥3 systemic reactogenicity symptoms. The adjusted mean (95% CI) number of systemic symptoms was 1.8 (1.6–2.0) and 3.2 (3.0–3.4), respectively. Local reactogenicity events (adjusted) were reported in 73.4% and 91.7% of participants who received NVX-CoV2373 and mRNA vaccines, respectively; the adjusted mean (95% CI) number of local symptoms was 1.5 (1.33–1.61) and 2.4 (2.31–2.52), respectively. These results support the use of adjuvanted, protein-based NVX-CoV2373 as an immunization option with low reactogenicity.

## 1. Introduction

To date, multiple types of vaccines have been developed to protect against COVID-19. As the need for additional COVID-19 vaccination continues, messenger RNA (mRNA)–based and protein-based vaccines are expected to be the most widely utilized platforms. The mRNA COVID-19 vaccines developed by Pfizer (BNT162b2) and Moderna (mRNA-1273) are approved for use in the United States (US) and Canada [1-3], and a protein-based vaccine formulated with saponin-based Matrix-M™ adjuvant developed by Novavax is authorized for use in the US and approved for use in Canada [3,4].

Participants in COVID-19 vaccine clinical trials have commonly reported mild and transient reactogenicity events within 7 days of receiving a vaccine, with many events resolving within 2 days post-vaccination [5-11]. These symptoms include local (injection-site) reactions (pain, tenderness, erythema, or swelling) or systemic reactions (fatigue, malaise, muscle pain, joint pain, nausea/vomiting, headache, or fever), which may increase with subsequent doses of a COVID-19 vaccine [6,9-12]. Local and systemic reactions are also commonly reported for immunizations targeting infectious diseases other than COVID-19 (e.g., influenza or shingles) [13,14]. Of note, studies comparing reactogenicity have generally found a higher incidence of local and systemic events following receipt of mRNA COVID-19 vaccines compared with influenza and other non–COVID-19 vaccines, as well as a higher frequency of reactogenicity-associated medication use, sick leave, and doctor’s visits [15,16].

COVID-19 vaccine–related reactogenicity events can affect work and other daily activities, leading to absenteeism from work [17,18] and presenteeism [17,19], as well as vaccine hesitancy [20-24]. Indeed, concern over vaccine side effects was found to be the most common reason for refusing an updated COVID-19 vaccine [24]. The 2019nCoV-406 study surveyed participants in the US and Canada who were receiving a COVID-19 vaccine to compare how reactogenicity impacted work and other daily activities [17]. Findings suggest that in the 6 days following vaccination, recipients of the protein-based NVX-CoV2373 vaccine had lower unadjusted reactogenicity rates and had trended toward less overall impairment relative to recipients of mRNA vaccines (BNT162b2 and mRNA-1273). We also observed that over 90% of the most frequently occurring solicited reactogenicity events were reported within the first 2 days after vaccination. Here, we present an additional analysis of the 2019nCoV-406 study, including adjustment for potential confounding, to more closely examine reactogenicity within 2 days of receiving an authorized/approved COVID-19 vaccine in previously vaccinated participants.

## 2. Materials and Methods

### 2.1 Study Design and Participants

The prospective, noninterventional, observational 2019nCoV-406 study investigated the impact of common reactogenicity events from the COVID-19 vaccine on absenteeism, presenteeism, and work productivity loss [17]. Briefly, the study enrolled working adults aged 18 to <65 years in the US and Canada who voluntarily received an authorized/approved primary series or booster (post–primary series) dose of a COVID-19 vaccine. As this was an observational study, participants selected the COVID-19 vaccine type they wanted to receive. A booster dose was defined as any COVID-19 dose received after completion of a primary COVID-19 vaccination series, regardless of prior vaccine type used or prior COVID-19 disease status. Post–primary series vaccinations included NVX-CoV2373 (5 μg recombinant spike protein co-formulated with 50 μg of Matrix-M adjuvant) or an mRNA vaccine (BTN162b2 [30 μg] and mRNA-1273 [50 μg]). Participants completed baseline/screening questionnaires on the day of their vaccine (day 0) and a daily diary for the following 6 days that included a Vaccine Symptoms Diary. Further details on participant inclusion criteria and study methods are available in the prior publication [17]. Written informed consent was provided by each participant prior to receipt of their requested vaccine dose and survey completion.

### 2.2 Objectives

The descriptive/comparative goal of this post hoc analysis was to determine the difference in local or systemic reactogenicity events occurring within the first 2 days after receipt of an NVX-CoV2373 vaccine versus an mRNA COVID-19 vaccine, administered after completion of any primary vaccination series.

### 2.3 Assessments

The Vaccine Symptoms Diary measured 11 solicited, participant-reported local/systemic symptoms over a 24-h recall period. Solicited systemic reactogenicity symptoms were fever, fatigue, malaise, muscle pain, joint pain, nausea/vomiting, and headache. Solicited local reactogenicity symptoms included pain, tenderness, swelling, and redness at the injection site. All symptoms were recorded using a 0-to-3 response scale based on the worst level of reactogenicity severity experienced, with responses categorized as 0 (no symptom present), 1 (mild/no interference with activity), 2 (moderate/interferes with activity), and 3 (severe/significant interference with activity). A participant was considered to have experienced a reactogenicity symptom if they reported a severity of at least grade 1. This analysis used data from the 2 days immediately following vaccination.

### 2.4 Statistics

Analyses of reactogenicity in the 2 days following vaccination were completed in the booster dose population (hereafter referred to as the post–primary series population), which consisted of participants who received any post–primary series dose of an approved/authorized COVID-19 vaccine, regardless of which vaccine type was used for prior doses or if they had previously had COVID-19. Due to low numbers receiving other vaccine types, only data related to NVX-CoV2373 or the mRNA vaccines (BNT162b2 and mRNA-1273) were included in this analysis. Estimation of the sample size required to power the primary objectives in the main study was described previously [17].

Results are presented by vaccine groups composed of participants who received NVX-CoV2373 and participants who received BNT162b2 or mRNA-1273, referred to as the mRNA vaccine group. Findings are also presented in subgroups for the different mRNA vaccines (BNT162b2 or mRNA-1273) and country (US or Canada).

Analyses are presented separately for systemic and local reactogenicity. The proportions of participants with any systemic/local reactogenicity and with individual systemic/local reactogenicity events, mean numbers of systemic/local events, and the proportion of participants with ≥3 systemic events are presented as comparative analyses. Comparative analyses were adjusted to address potential confounding using inverse probability of treatment weighting (IPTW). As described previously [17], each participant was assigned a propensity score based on a select group of demographic and clinical characteristics identified using standardized differences. The present analysis used the following covariates in the IPTW model: country (US vs Canada), prior COVID-19 diagnosis (yes vs no), race (Asian, White), job category (professional), work at home (yes, no, or prefer not to answer), gender identity (male, female), and scheduled to work in the next 24 h (yes vs no). The comparative analyses were weighted using stabilized inverse probability of treatment weights. Because of the sample size limitation, event severity and all subgroup analyses were analyzed descriptively. All results use the overall participant post–primary series sample. Results were not adjusted for multiple comparisons.

## 3. Results

### 3.1 Participants

The 2019nCoV-406 study was conducted between July 2022 and March 2023. The post–primary series population included 1130 participants, 303 of whom received NVX-CoV2373 and 827 who received an mRNA vaccine during the study period. Baseline demographics and clinical characteristics were generally balanced between vaccine groups, as reported previously [17]; however, some differences between the NVX-CoV2373 and mRNA vaccine groups were observed related to ethnicity (**Table 1**). A higher proportion of Hispanic, Latin American, or Latinx (50.8%) and White (50.2%) participants received NVX-CoV2373 versus an mRNA vaccine (25.0% and 33.6%, respectively). By contrast, more Asian (22.9%) and Native Hawaiian or Pacific Islander (9.1%) participants received an mRNA vaccine versus NVX-CoV2373 (13.2% and 2.0%, respectively). Medical conditions that put participants at high risk for severe COVID-19 were relatively low in both groups (NVX-CoV2373, 6.3%; mRNA, 5.4%). Of those participants who received NVX-CoV2373, more did so as a first (60.7%) versus second (39.3%) dose after completion of the primary series. Alternatively, of those who received an mRNA vaccine, more received this as a second (62.6%) versus first (37.4%) post–primary series dose.

**Table 1.** Baseline demographics and clinical characteristics of study participants by region.

| Parameter | Post–primary series<br>population [17] |  | US |  | Canada |  |
| --- | --- | --- | --- | --- | --- | --- |
|  | NVX-CoV2373 | mRNA | NVX-CoV2373 | mRNA | NVX-CoV2373 | mRNA |
|  | (n = 303) | vaccine<sup>a</sup><br>(n = 827) | (n = 205) | vaccine<sup>a</sup><br>(n = 426) | (n = 98) | vaccine<sup>a</sup><br>(n = 401) |
| **Age, mean (SD) years** | 38.9 (11.8) | 40.1 (13.0) | 39.4 (12.0) | 42.6 (13.4) | 37.9 (11.2) | 37.5 (12.0) |
| **Gender identity, n (%)** |  |  |  |  |  |  |
| Female | 156 (51.5) | 469 (56.7) | 109 (53.2) | 243 (57.0) | 47 (48.0) | 226 (56.4) |
| Male | 142 (46.9) | 355 (42.9) | 95 (46.3) | 182 (42.7) | 47 (48.0) | 173 (43.1) |
| Genderfluid | 1 (0.3) | 0 | 0 | 0 | 1 (1.0) | 0 |
| Nonbinary | 2 (0.7) | 3 (0.4) | 0 | 1 (0.2) | 2 (2.0) | 2 (0.5) |
| Prefer not to answer | 2 (0.7) | 0 | 1 (0.5) | 0 | 1 (1.0) | 0 |
| **Race/ethnicity<sup>b</sup>, n (%)** |  |  |  |  |  |  |
| African American or Black | 33 (10.9) | 77 (9.3) | 27 (13.2) | 67 (15.7) | 6 (6.1) | 10 (2.5) |
| Asian<sup>c</sup> | 40 (13.2) | 189 (22.9) | 8 (3.9) | 21 (4.9) | 32 (32.7) | 168 (41.9) |
| Hispanic, Latin American, or Latinx | 154 (50.8) | 207 (25.0) | 145 (70.7) | 197 (46.2) | 9 (9.2) | 10 (2.5) |
| Middle Eastern or North African<sup>d</sup> | 5 (1.7) | 21 (2.5) | 2 (1.0) | 2 (0.5) | 3 (3.1) | 19 (4.7) |
| Native Hawaiian or Pacific Islander<sup>e</sup> | 6 (2.0) | 75 (9.1) | 2 (1.0) | 0 | 4 (4.1) | 75 (18.7) |
| White | 152 (50.2) | 278 (33.6) | 106 (51.7) | 157 (36.9) | 46 (46.9) | 121 (30.2) |
| Other<sup>f</sup> | 11 (3.6) | 22 (2.7) | 6 (2.9) | 9 (2.1) | 5 (5.1) | 13 (3.2) |
| **Prior COVID-19 diagnosis, n (%)** | 119 (39.3) | 433 (52.4) | 67 (32.7) | 239 (56.1) | 52 (53.1) | 194 (48.4) |
| **Medical condition that puts participant at high risk for severe COVID-19<sup>b</sup>, n (%)** |  |  |  |  |  |  |
| Diabetes | 6 (31.6) | 21 (46.7) | 2 (1.0) | 15 (3.5) | 4 (4.1) | 6 (1.5) |
| Hypertension | 7 (36.8) | 13 (28.9) | 5 (2.4) | 10 (2.3) | 2 (2.0) | 3 (7.5) |
| Heart disease | 2 (10.5) | 8 (17.8) | 1 (6.7) | 8 (1.9) | 1 (1.0) | 0 |
| Respiratory conditions | 5 (26.3) | 11 (24.4) | 4 (2.0) | 7 (1.6) | 1 (1.0) | 4 (1.0) |
| Other | 5 (26.3) | 13 (28.9) | 4 (2.0) | 10 (2.3) | 1 (1.0) | 3 (0.7) |
| **Booster dose, n (%)** |  |  |  |  |  |  |
| First | 184 (60.7) | 309 (37.4) | 167 (81.5) | 266 (62.4) | 17 (17.3) | 43 (10.7) |
| Second or later | 119 (39.3) | 518 (62.6) | 38 (18.5) | 160 (37.6) | 81 (82.7) | 358 (89.3) |
| **mRNA vaccine type, n (%)** |  |  |  |  |  |  |
| Monovalent | - | 652 (78.8) | - | 401 (94.1) | - | 251 (62.6) |
| Bivalent | - | 175 (21.2) | - | 25 (5.9) | - | 150 (37.4) |

Baseline demographics and clinical characteristics for the post–primary series population have been published previously [17]. SD = standard deviation. <sup>a</sup>Individuals received either BNT162b2 or mRNA-1273. <sup>b</sup>The categories for these variables were not mutually exclusive (participants could have listed more than one). <sup>c</sup>Includes participants who identified as Chinese, South Asian (e.g., East Indian, Pakistani, or Sri Lankan), Southeast Asian (e.g., Vietnamese, Cambodian, Laotian, or Thai), Korean, or Japanese. <sup>d</sup>Includes participants who identified as Middle Eastern, North African, Arab, or West Asian (e.g., Iranian or Afghan). <sup>e</sup>Includes participants who identified as Native Hawaiian, Pacific Islander, or Filipino. <sup>f</sup>Race/ethnicity categories with fewer than 20 responses are captured in the “other” category and include: Alaska Native, American Indian, or Native American participants (total n = 6); race or ethnicity not listed (n = 16); and prefer not to answer (n = 11).

Of the 1130 participants in the post–primary series population, 631 participants (NVX-CoV2373, n = 205; mRNA, n = 426) were from the US and 499 (NVX-CoV2373, n = 98; mRNA, n = 401) were from Canada (**Table 1**). Baseline demographics and characteristics of participants from the US and Canada were generally similar to those observed in the overall post–primary series population. Most participants with a Hispanic, Latin American, or Latinx ethnicity came from the US and most participants with an Asian or Native Hawaiian or Pacific Islander ethnicity came from Canada.

### 3.2 Systemic Reactogenicity

Within the first 2 days after vaccination, solicited systemic reactogenicity events were reported in 56.4% of participants who received NVX-CoV2373 and 84.4% of participants who received an mRNA vaccine (BNT162b2: 84.5%; mRNA-1273: 84.3%). After IPTW adjustment, 60.5% of participants who received the NVX-CoV2373 reported solicited systemic reactogenicity events compared with 83.8% of participants who received an mRNA vaccine (**Figure 1**). Muscle pain (NVX-CoV2373: 41.6%; mRNA vaccine: 72.3%), fatigue (47.8% and 66.5%, respectively), and malaise (34.5% and 57.7%, respectively) were the most common systemic events in each vaccine group.

**Figure 1.**
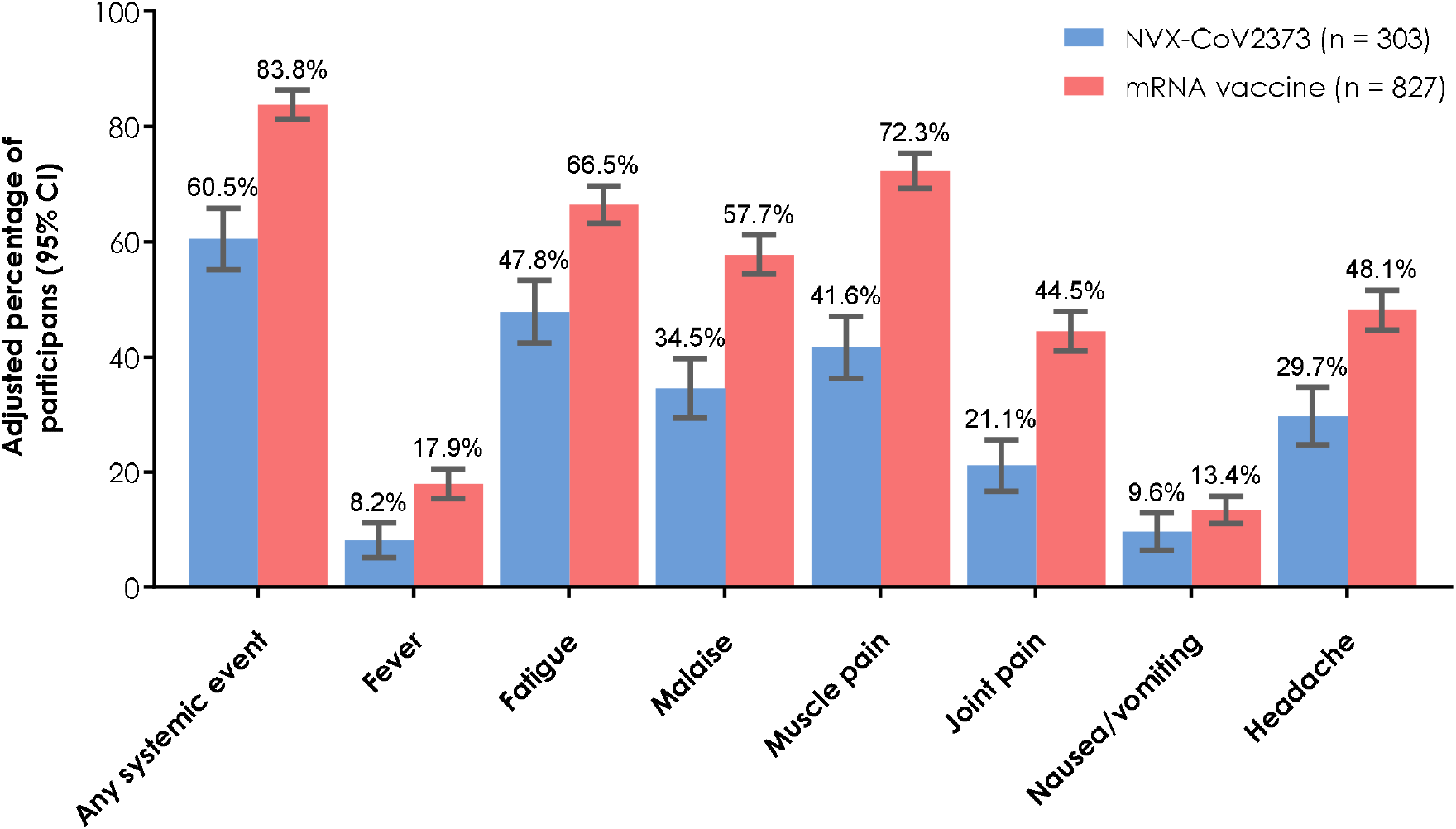
Rates of overall and individual solicited systemic reactogenicity events within 2 days of post–primary-series vaccination (IPTW adjusted estimates). Results are presented for the post–primary series population. CI, confidence interval; IPTW, inverse probability of treatment weighting.

Participants who received NVX-CoV2373 reported a mean number (SD) of 1.8 (2.0) systemic events, whereas those who received an mRNA vaccine reported a mean of 3.2 (2.1) systemic events (**Table 2**). Adjusting for confounding by IPTW led to similar results, with mean numbers of events (95% CI) of 1.8 (1.6–2.0) for the NVX-CoV2373 group and 3.2 (3.0–3.4) for the mRNA vaccine group (**Figure 2**). Markedly fewer participants who received NVX-CoV2373 reported three or more systemic reactogenicity events than those who received an mRNA vaccine (adjusted values [95% CI]: NVX-CoV2373, 33.9% [28.7–39.1%]; mRNA vaccine, 61.4% [58.1–64.8%]).

**Table 2.** Descriptive analysis of systemic reactogenicity events in the overall population and by mRNA vaccine subgroup (unadjusted)

|  | NVX-CoV2373<br>(n = 303) | mRNA vaccine<br>(n = 827) | mRNA vaccine subgroup |  |
| --- | --- | --- | --- | --- |
|  |  |  | BNT162b2<br>(n = 502) | mRNA-1273<br>(n = 325) |
| Median (range) | 1 (0–7) | 3 (0–7) | 3 (0–7) | 4 (0–7) |
| Number of events, n (%) |  |  |  |  |
| No systemic reactogenicity events | 132 (43.6) | 129 (15.6) | 78 (15.5) | 51 (15.7) |
| 1 | 41 (13.5) | 91 (11.0) | 62 (12.4) | 29 (8.9) |
| 2 | 34 (11.2) | 98 (11.9) | 73 (14.5) | 25 (7.7) |
| 3 | 26 (8.6) | 108 (13.1) | 69 (13.7) | 39 (12.0) |
| 4 | 26 (8.6) | 140 (16.9) | 81 (16.1) | 59 (18.2) |
| 5 | 26 (8.6) | 128 (15.5) | 72 (14.3) | 56 (17.2) |
| 6 | 11 (3.6) | 89 (10.8) | 48 (9.6) | 41 (12.6) |
| 7 | 7 (2.3) | 44 (5.3) | 19 (3.8) | 25 (7.7) |
| Severity, n (%) |  |  |  |  |
| No reactogenicity symptoms reported | 132 (43.6) | 129 (15.6) | 78 (15.5) | 51 (15.7) |
| Mild/no interference with activities | 87 (28.7) | 290 (35.1) | 184 (36.7) | 106 (32.6) |
| Moderate/interfered with activities | 62 (20.5) | 288 (34.8) | 177 (35.3) | 111 (34.2) |
| Severe/significant interference with activities | 22 (7.3) | 120 (14.5) | 63 (12.5) | 57 (17.5) |

Table shows descriptive data summarizing systemic reactogenicity events reported within 2 days of receipt of a post-primary series vaccination in the post-primary series population. All values are unadjusted.

**Figure 2.**
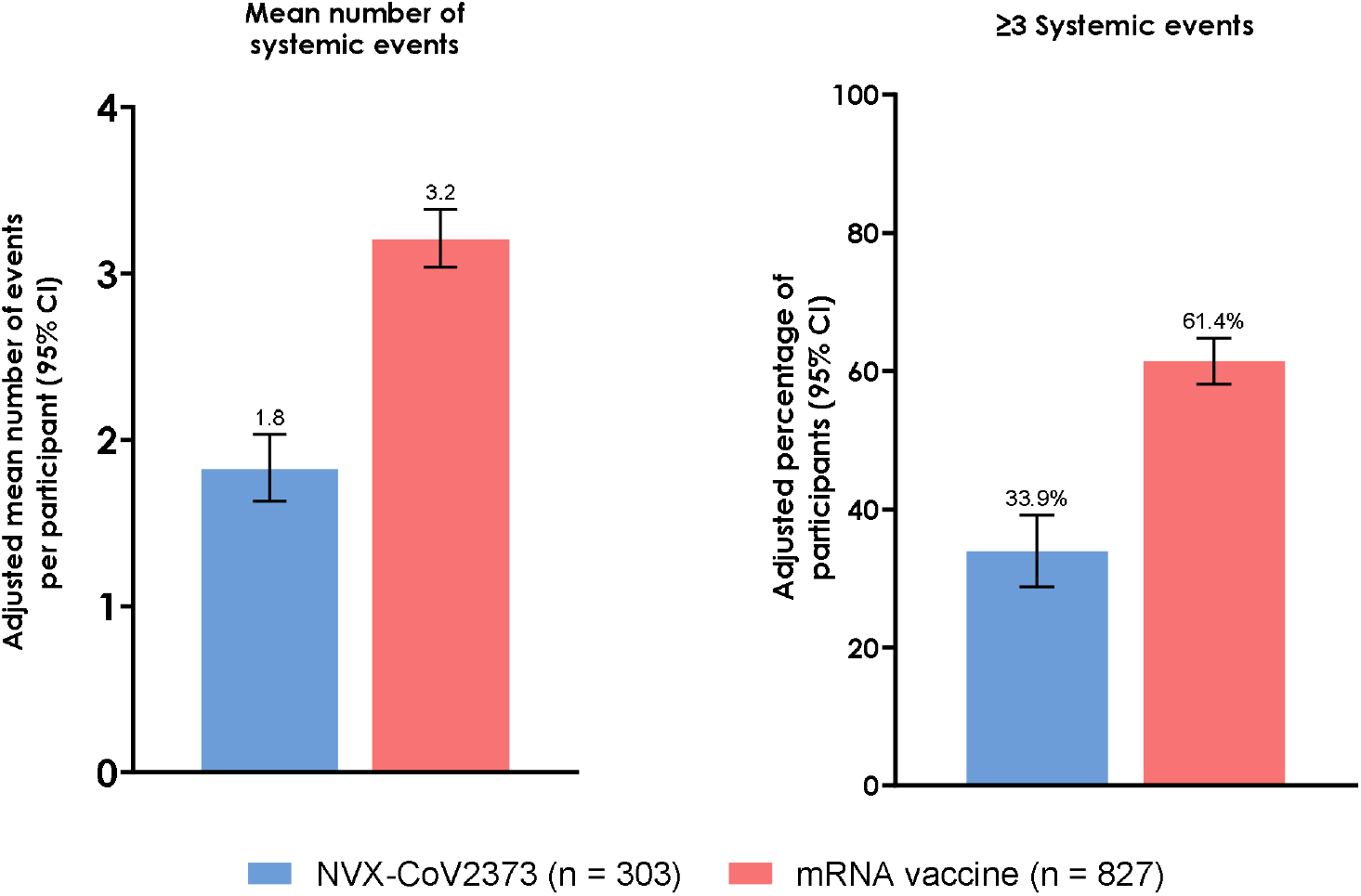
Summary of solicited systemic reactogenicity events within 2 days of post–primary series vaccination (IPTW adjusted estimates). Post–primary series population. CI, confidence interval; IPTW, inverse probability of treatment weighting.

With respect to event severity, a lower proportion (unadjusted) of participants who received NVX-CoV2373 reported a moderate or severe/significant systemic event within 2 days of vaccination compared with participants who received an mRNA vaccine (27.7% [84/303] vs 49.3% [408/827]). Mild events were also reported in lower proportions of participants who received NVX-CoV2373 (28.7%) compared with those who received an mRNA vaccine (35.1%).

The mean (SD) number of systemic events was similar whether participants received BNT162b2 (3.0 [2.1]) or mRNA-1273 (3.5 [2.2]). Similarly, the proportion of participants reporting any systemic event was 84.5% (424/502) and 84.3% (274/325), respectively. However, a higher proportion of participants receiving mRNA-1273 reported three or more systemic reactogenicity events (unadjusted; 67.7% [220/325]) compared with BNT162b2 (57.6% [289/502]). The proportion of participants reporting moderate-to-severe systemic events was 51.7% (168/325) and 47.8% (240/502) with mRNA-1273 and BNT162b2, respectively (**Table 2**).

When assessed by country, unadjusted proportions of systemic reactogenicity events in the US were lower in participants who received NVX-CoV2373 (48.5%) compared with participants who received an mRNA vaccine (79.1%) (**Table 3**). Corresponding proportions tended to be higher for both vaccine types in Canadian participants (NVX-CoV2373: 72.5%; mRNA: 90.0%). Regardless of country, participants who received NVX-CoV2373 reported fewer systemic reactogenicity events (mean [SD]: US, 1.6 [2.1]; Canada, 2.1 [2.0]) than those who received an mRNA vaccine (mean [SD]: US, 3.1 [2.3]; Canada, 3.3 [1.9]). Similarly, regardless of country, fewer participants who received NVX-CoV2373 reported three or more events (US: 29.8% [61/205]; Canada: 35.7% [35/98]) compared with those who received an mRNA vaccine (US: 58.2% [248/426]; Canada: 64.8% [260/401]) and fewer participants who received NVXCoV2373 reported moderate-to-severe events (US: 22.0% [45/205]); Canada: 39.8% [39/98]) compared with those who received an mRNA vaccine (US: 45.5% [194/426]; Canada: 53.4% [214/401]).

**Table 3.** Descriptive analysis of systemic reactogenicity events by country (unadjusted)

|  | US |  | Canada |  |
| --- | --- | --- | --- | --- |
|  | NVX-CoV2373<br>(n = 205) | mRNA vaccine<br>(n = 426) | NVX-CoV2373<br>(n = 98) | mRNA vaccine<br>(n = 401) |
| Mean (SD) | 1.6 (2.1) | 3.1 (2.3) | 2.1 (2.0) | 3.3 (1.9) |
| Median (range) | 0 (0–7) | 3 (0–7) | 2 (0–7) | 4 (0–7) |
| Any systemic event, n (%) | 100 (48.5) | 337 (79.1) | 71 (72.5) | 361 (90.0) |
| Number of events, n (%) |  |  |  |  |
| No systemic reactogenicity events | 105 (51.2) | 89 (20.9) | 27 (27.6) | 40 (10.0) |
| 1 | 23 (11.2) | 41 (9.6) | 18 (18.4) | 50 (12.5) |
| 2 | 16 (7.8) | 48 (11.3) | 18 (13.4) | 50 (12.5) |
| 3 | 18 (8.8) | 52 (12.2) | 8 (8.2) | 56 (14.0) |
| 4 | 12 (5.9) | 57 (13.4) | 14 (14.3) | 83 (20.7) |
| 5 | 20 (9.8) | 63 (14.8) | 6 (6.1) | 65 (16.2) |
| 6 | 6 (2.9) | 45 (10.6) | 5 (5.1) | 44 (11.0) |
| 7 | 5 (2.4) | 31 (7.3) | 2 (2.0) | 12 (3.2) |
| Severity, n (%) |  |  |  |  |
| No reactogenicity symptoms reported | 105 (51.2) | 89 (20.9) | 27 (27.6) | 40 (10.0) |
| Mild/no interference with activities | 55 (26.8) | 143 (33.6) | 32 (32.7) | 147 (36.7) |
| Moderate/interfered with activities | 31 (15.1) | 139 (32.5) | 31 (31.6) | 149 (37.2) |
| Severe/significant interference with activities | 14 (6.8) | 55 (12.9) | 8 (8.2) | 65 (16.2) |

Table shows descriptive data summarizing systemic reactogenicity events reported within 2 days of receipt of a post–primary series vaccination in the post–primary series population. All values are unadjusted.

### 3.3 Local Reactogenicity

Similar to systemic reactogenicity, a lower proportion of participants who received NVX-CoV2373 (68.3%) experienced at least one solicited local reactogenicity event, compared with those who received an mRNA vaccine (91.9%; BNT162b2, 92.6%; mRNA-1273, 90.8%). After IPTW adjustment, local reactogenicity events were estimated to be reported in 73.7% and 91.7% of participants who received a booster dose of NVX-CoV2373 and mRNA vaccine, respectively (**Figure 3**). This trend in differences continued, with the overall frequency of each individual event occurring in a higher proportion of participants in the mRNA vaccine versus NVX-CoV2373 group. For both the NVX-CoV2373 and mRNA vaccine groups, the most common solicited local events were pain (61.7% and 84.8%, respectively) and tenderness (65.4% and 87.9%, respectively) at the injection site.

**Figure 3.**
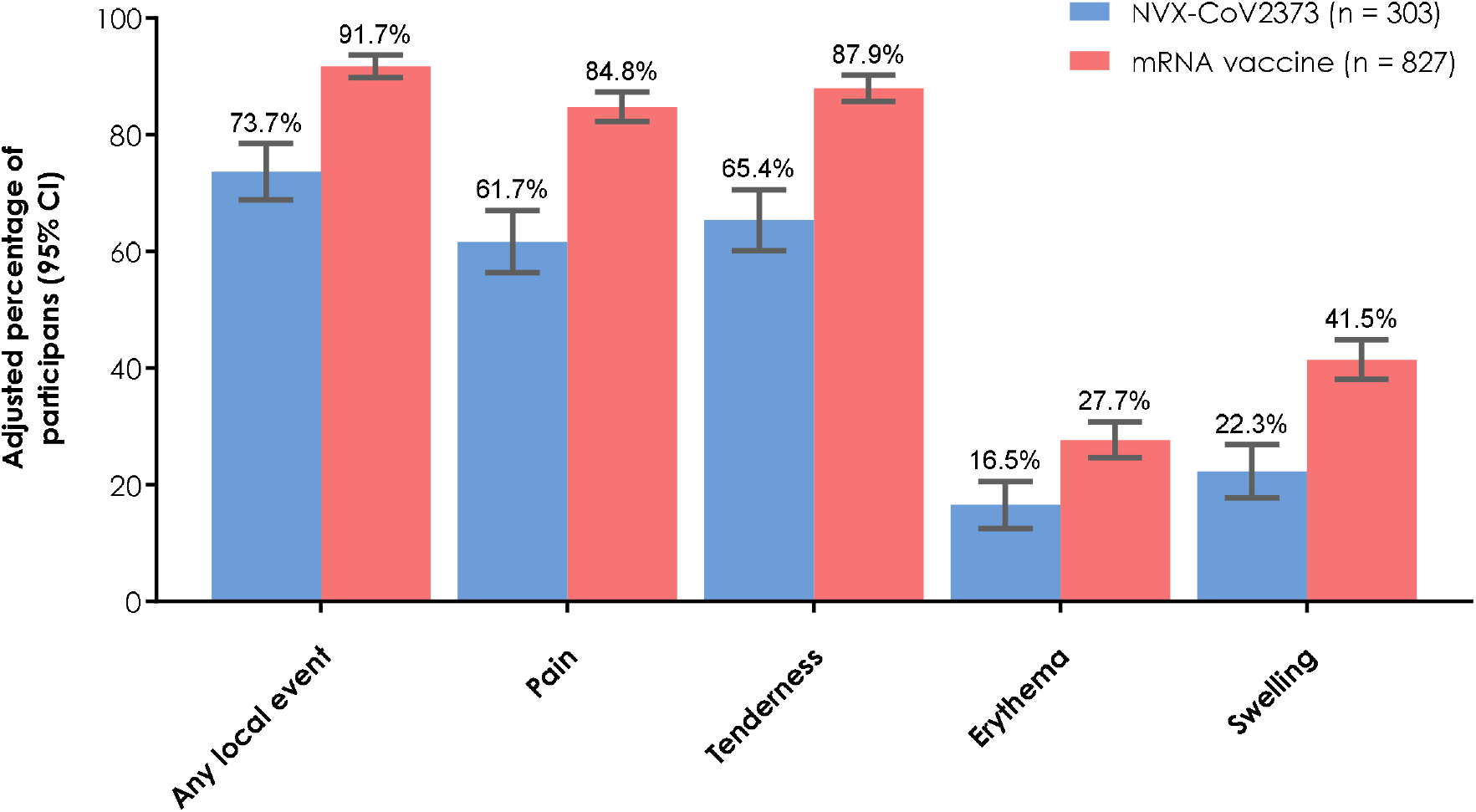
Overall and individual solicited local reactogenicity events within 2 days of post–primary series vaccination (IPTW adjusted estimates). Post–primary series population. CI, confidence interval; IPTW, inverse probability of treatment weighting.

The mean number (SD) of reported local reactogenicity events per individual was 1.5 (1.3) for participants who received NVX-CoV2373 and 2.4 (1.1) for those who received an mRNA vaccine. Mean (95% CI) adjusted numbers were 1.5 (1.3–1.6) and 2.4 (2.3–2.5), respectively. Compared with the NVX-CoV2373 group (21.1%), the proportion (unadjusted) of participants reporting moderate or severe/significant local reactogenicity events was 2.5-fold higher for participants in the mRNA vaccine group (52.0%) (**Table 4**).

**Table 4.** Descriptive analysis of local reactogenicity events in the overall population and by mRNA vaccine subgroup (unadjusted)

|  | NVX-CoV2373<br>(n = 303) | mRNA vaccine<br>(n = 827) | mRNA vaccine subgroup |  |
| --- | --- | --- | --- | --- |
|  |  |  | BNT162b2<br>(n = 502) | mRNA-1273<br>(n = 325) |
| Median (range) | 2 (0–4) | 2 (0–4) | 2 (0–4) | 3 (0–4) |
| Number of events, n (%) |  |  |  |  |
| No systemic reactogenicity events | 96 (31.7) | 67 (8.1) | 37 (7.4) | 30 (9.2) |
| 1 | 51 (16.8) | 57 (6.9) | 38 (7.6) | 19 (5.8) |
| 2 | 98 (32.3) | 328 (39.7) | 227 (45.2) | 101 (31.1) |
| 3 | 33 (10.9) | 216 (26.1) | 130 (25.9) | 86 (26.5) |
| 4 | 25 (8.3) | 159 (19.2) | 70 (13.9) | 89 (27.4) |
| Severity, n (%) |  |  |  |  |
| No reactogenicity symptoms reported | 96 (31.7) | 67 (8.1) | 37 (7.4) | 30 (9.2) |
| Mild/no interference with activities | 143 (47.2) | 330 (39.9) | 222 (44.2) | 108 (33.2) |
| Moderate/interfered with activities | 57 (18.8) | 338 (40.9) | 200 (39.8) | 138 (42.5) |
| Severe/significant interference with activities | 7 (2.3) | 92 (11.1) | 43 (8.6) | 49 (15.1) |

Table shows descriptive data summarizing local reactogenicity events reported within 2 days of receipt of a post–primary series vaccination in the post–primary series population. All values are unadjusted.

Reporting of local events was generally similar whether participants received BNT162b2 or mRNA-1273 (**Table 4**). The proportion (unadjusted) of participants reporting any local event was 92.6% (465/502) for BNT162b2 and 90.8% (295/325) for mRNA-1273. The mean number (SD) of local events reported was similar between subgroups (2.3 [1.0] and 2.6 [1.2], respectively); however, the proportion of participants who reported moderate or severe local events was higher in recipients of mRNA-1273 (57.5% [187/325]) than recipients of BNT162b2 (48.4% [243/502]).

In both the US and Canada, proportions (unadjusted) of local reactogenicity events were lower in participants who received NVX-CoV2373 compared with participants who received an mRNA vaccine, with events reported at a lower frequency in US versus Canadian participants (US: NVX-CoV2373, 62.4% vs mRNA, 88.3%; Canada: NVX-CoV2373, 80.6% vs mRNA, 95.8%) (**Table 5**). The mean number of local reactogenicity events reported in the US and Canada tended to follow the same pattern as the overall post–primary series population, with fewer events reported for those who received NVX-CoV2373 versus an mRNA vaccine. Similar to the overall population, regardless of country, lower proportions of participants who received NVX-CoV2373 reported moderate-to-severe local reactogenicity events (US: 20.0% [41/205]; Canada: 23.5% [23/98]) compared with those who received an mRNA vaccine (US: 46.0% [196/426]; Canada: 58.4% [234/401]).

**Table 5.** Descriptive analysis of local reactogenicity events by country (unadjusted)

|  | US |  | Canada |  |
| --- | --- | --- | --- | --- |
|  | NVX-CoV2373<br>(n = 205) | mRNA vaccine<br>(n = 426) | NVX-CoV2373<br>(n = 98) | mRNA vaccine<br>(n = 401) |
| Mean (SD) | 1.3 (1.3) | 2.4 (1.3) | 1.7 (1.2) | 2.4 (1.0) |
| Median (range) | 1 (0–4) | 2 (0–4) | 2 (0–4) | 2 (0–4) |
| Any local event, n (%) | 128 (62.4) | 376 (88.3) | 79 (80.6) | 384 (95.8) |
| Number of events, n (%) |  |  |  |  |
| No local reactogenicity events | 77 (37.6) | 50 (11.7) | 19 (19.4) | 17 (4.2) |
| 1 | 33 (16.1) | 36 (8.5) | 18 (18.4) | 21 (5.2) |
| 2 | 59 (28.8) | 133 (31.2) | 39 (39.8) | 195 (48.6) |
| 3 | 20 (9.8) | 108 (25.4) | 12 (13.3) | 108 (26.9) |
| 4 | 16 (7.8) | 99 (23.2) | 9 (9.2) | 60 (15.0) |
| Severity, n (%) |  |  |  |  |
| No reactogenicity symptoms reported | 77 (37.6) | 50 (11.7) | 19 (19.4) | 17 (4.2) |
| Mild/no interference with activities | 87 (42.4) | 180 (42.3) | 56 (57.1) | 150 (37.4) |
| Moderate/interfered with activities | 37 (18.1) | 147 (34.5) | 20 (20.4) | 191 (47.6) |
| Severe/significant interferences with activities | 4 (2.0) | 49 (11.5) | 3 (3.1) | 43 (10.7) |

Table shows descriptive data summarizing local reactogenicity events reported within 2 days of receipt of a post–primary series vaccination in the post–primary series population. All values are unadjusted.

## 4. Discussion

In this post hoc analysis of the 2019nCoV-406 study, participants who received NVX-CoV2373 after completing a primary vaccination series reported fewer and less severe local and systemic reactogenicity symptoms in the 2 days following vaccination compared with those who received an mRNA vaccine after completing a primary vaccination series. These real-world data provide a better understanding of reactogenicity events associated with the two COVID-19 vaccine platforms (protein-based and mRNA) received in the US and Canada. In the 2 days following vaccination, local and systemic reactogenicity events were more commonly reported in participants who received an mRNA vaccine than in those who received NVX-CoV2373. The proportions of participants reporting the most common local (injection site pain and tenderness) and systemic (muscle pain, fatigue, and malaise) reactogenicity events were higher among those who received an mRNA vaccine (>84% and >57%, respectively) than among those who received NVX-CoV2373 (up to 65% and up to 48%, respectively). In addition, mRNA vaccine recipients were more likely to report experiencing three or more systemic events or events with moderate-to-severe severity than participants who received NVX-CoV2373. These findings are consistent regardless of the mRNA brand compared.

These data build on overall findings from the 2019nCoV-406 study [17] by focusing on the 2 days immediately following receipt of a post–primary series vaccination and adjusting for potential confounding using IPTW. As noted previously, this timing is of interest because over 90% of the most frequently occurring solicited reactogenicity events in the 2019nCoV-406 study were reported within the first 2 days. Similarly, multiple studies have shown that systemic and local reactogenicity events are short-lived, with mean or median durations of 2 days or less for the most frequently reported solicited reactogenicity events [7-9,11]. The primary analysis from the 2019nCoV-406 study suggested a trend toward less overall work impairment with NVX-CoV2373 compared with the mRNA vaccines [17]. In addition, greater proportions of participants who received an mRNA vaccine reported local and systemic reactogenicity events compared with NVX-CoV2373. This post hoc analysis showed consistent findings for the 2 days after post–primary series vaccination, including when potential confounding related to differences in baseline and clinical characteristics were accounted for using IPTW.

Multiple studies have found greater reactogenicity of mRNA vaccines relative to other COVID-19 vaccine platforms, including adjuvanted protein-based vaccines such as NVX-CoV2373 [12,16,25-30]. While there are limitations to direct comparisons between different studies, it is notable how closely these findings from the 2019nCoV-406 study mirror observations from the Oxford COV-BOOST trial and a National Institute for Allergy and Infectious Diseases and National Institutes of Health–funded booster study [26,27]. Results from this analysis of 2019nCoV-406 expand on previous reports [16,28,30] by focusing on the effects of vaccine doses administered after completion of primary COVID-19 vaccination series. Additional COVID-19 vaccine dosing, whether as the second dose of the primary series or as subsequent doses, can result in greater reactogenicity compared with the first injection [6,9,10], especially when receiving a heterologous booster [19,25,31]. Although the studies investigating reactogenicity in homologous and heterologous booster doses were generally not powered to make comparisons between vaccine types, altogether findings from 2019nCoV-406 and other studies suggest that NVX-CoV2373, when administered as a heterologous booster, has less reactogenicity than mRNA vaccines when administered as a homologous or heterologous post–primary series dose [12,25,26,30]. This is especially relevant as most people in the US and Canada have received a primary series of an mRNA vaccine. Concern about side effects is a primary reason for COVID-19 vaccine hesitancy [22-24]. Accordingly, COVID-19 vaccines with lower rates of reactogenicity, which is suggested of heterologous use of NVX-CoV2373 by the present analysis and several descriptive studies [12,25,26,30], have the potential to decrease vaccine hesitancy. This may be especially useful given the continued evolution of SARS-CoV-2 and recommendations from public health authorities for annual updates to the COVID-19 vaccine strain composition [32-35], suggesting the potential need for seasonal COVID-19 vaccination, similar to influenza vaccination.

Comparison between the two mRNA vaccines showed that BNT162b2 (30 μg) tended to elicit fewer and slightly less severe systemic and local reactogenicity events than mRNA-1273 (50 μg). If making cross-study comparisons, it is important to note the vaccine doses investigated in the studies [12,25-28]. In particular, participants in COV-BOOST and the MixNMatch Study (DMID 21-0012) received a higher dose of mRNA-1273 (100 μg) than is currently approved [26,27]. However, mRNA-1273 was administered at the approved 50 μg dose [2] in the 2019nCoV-406 study, and rates of reactogenicity events observed with RNA-1273 were still higher than those observed with BNT162b2.

Limitations to the 2019nCoV-406 study have been previously reported [17] and are inherent to all noninterventional, real-world investigations. For example, study participants may not be representative of all populations receiving COVID-19 vaccines. This may be due to the timing of the study relative to vaccine approval/authorization; the later authorization for use and availability of NVX-CoV2373 (winter/early spring 2023) relative to that of mRNA vaccines (late summer/early fall 2022) may have created a temporal bias. Low enrollment at some study sites likely led to the concentration of participants with particular demographics (e.g., race/ethnicity) or with access to a specific type of vaccine. In addition, while availability of different vaccine types may have been defined by what was available at the site, vaccines were selected by the participant, which could also introduce bias. Real-word studies using patient-reported outcomes are often associated with concerns regarding the completeness and accuracy of patient reporting. While participants were instructed to complete the daily questions at the same time every day, it was possible to access and complete the diaries within an 8-h period.

It is relevant to note that the study was not powered to evaluate reactogenicity and that the post hoc nature of this analysis limits the conclusions that can be derived from the results. IPTW adjustments were made for the overall post-primary dose population; however, adjusted analyses were not available for data reporting on severity or for the mRNA vaccine and country subgroups. Results in the country subgroups were largely similar to those of the overall population, however some differences were observed. A higher proportion of participants in the US (32% [205/630]) selected NVX-CoV2373 compared with participants in Canada (20% [98/499]); reactogenicity tended to be reported in higher proportions of Canadian participants, regardless of the vaccine received, and the differences in reactogenicity reporting between NVX-CoV2373 and mRNA vaccine subgroups tended to be greater in participants in the US compared with those from Canada. Some of these findings may be limited by differences in adverse event reporting in the US and Canada; however, reactogenicity was higher among recipients of mRNA vaccines compared with NVX-CoV2373 in both countries. Finally, the impact of post–primary series dose number (e.g., first vs second post–primary series dose) relative to reactogenicity was not assessed.

The 2019nCoV-406 study provides data from a large, real-world population of participants receiving an additional dose of COVID-19 vaccine after completion of a primary vaccination series. Reactogenicity was captured in the same manner as in the phase 3 studies of COVID-19 vaccines, thereby strengthening the generalizability of these findings. Importantly, participants were enrolled at the time of their vaccination, with data collected daily, improving accuracy and limiting recall bias. In addition, this post hoc analysis used propensity score adjustments (i.e., IPTW) to reduce bias.

## 5. Conclusions

Considering the lower frequency and intensity of COVID-19 reactogenicity symptoms observed in this post hoc analysis of the real-world 2019nCoV-406 study, this analysis supports use of adjuvanted protein-based NVX-CoV2373 as an immunization option that has low reactogenicity. Future prospective studies are needed to confirm the observations reported here and to continue to examine the effects of age, sex, race/ethnicity, and even comorbidities on COVID-19 vaccine reactogenicity.

## Data Availability

Data are available on request due to privacy restrictions. The data presented in this study are available in aggregate form on request to the corresponding author. The data are not publicly available due to privacy requirements.

## Author Contributions

Conceptualization, K.H., D.O., H.B., and S.T.; methodology, M.D.R., K.H., R.Z., D.O., S.O., L.J., and S.T.; soft-ware, R.Z., S.O., and L.J.; validation, R.Z., D.O., S.O. and L.J.; formal analysis, M.D.R., K.H., R.Z., and D.O.; investigation, all authors; resources, M.D.R., A.M.M., M.M., H.B., and S.T.; data curation, R.Z., S.O., and L.J.; writing—original draft preparation, M.D.R., K.H., R.Z., D.O., S.O., L.J., and S.T.; writing—review and editing, all authors; visualization, R.Z., D.O., and A.M.M.; supervision, M.D.R. and S.T.; project administration, S.O. and L.J.; funding acquisition, M.D.R. and S.T. All authors have read and agreed to the published version of this manuscript.

## Funding

This research was funded by Novavax, Inc., and the APC was funded by Novavax, Inc.

## Institutional Review Board Statement

This study was conducted in accordance with the Declaration of Helsinki and approved by the Institutional Review Board (or Ethics Committee) of Advarra (Approval notice MOD01579170, approved 24 February 2023).

## Informed Consent Statement

Informed consent was obtained from all subjects involved in this study.

## Acknowledgments

Medical writing and editorial support were provided by Miranda Bader-Goodman, PhD, Meredith Kalish, MD, CMPP, Kelly M. Fahrbach, PhD, CMPP, and Ebenezer M. Awuah-Yeboah, BS, of Ashfield MedComms, an Inizio company, and funded by Novavax, Inc. The authors would like to thank Angela Miller for her operational involvement in the study.

## Conflicts of Interest

MDR, AMM, ST, HB, and MM are employees of Novavax, Inc. and may hold stock in Novavax, Inc. KH, RZ, SO, DO, and LJ are employees of RTI Health Solutions, an independent research consultancy that received funding under a research contract with Novavax, Inc. to conduct this study. The funder participated in the design and interpretation of the data and the writing of this manuscript but had no role in the collection or analysis of the data or in the decision to publish these results.

## Legal Disclosure

Registered and proprietary names, e.g., trademarks, used herein are not to be considered unprotected by law even when not specifically marked as such.

## References

1. Food and Drug Administration. Comirnaty. Available online: https://www.fda.gov/vaccines-blood-biologics/comirnaty (accessed on 23 May 2024).

2. Food and Drug Administration. Spikevax. Available online: https://www.fda.gov/vaccines-blood-biologics/spikevax (accessed on 24 May 2024).

3. Health Canada. Approved COVID-19 Vaccines. Available online: https://www.canada.ca/en/health-canada/services/drugs-health-products/covid19-industry/drugs-vaccines-treatments/vaccines.html (accessed on 30 May 2024).

4. Food and Drug Administration. Novavax COVID-19 Vaccine, Adjuvanted. Available online: https://www.fda.gov/vaccines-blood-biologics/coronavirus-covid-19-cber-regulated-biologics/novavax-covid-19-vaccine-adjuvanted (accessed on 24 May 2024).

5. Anez, G.; Dunkle, L.M.; Gay, C.L.; Kotloff, K.L.; Adelglass, J.M.; Essink, B.; Campbell, J.D.; Cloney-Clark, S.; Zhu, M.; Plested, J.S.; et al. Safety, Immunogenicity, and efficacy of the NVX-CoV2373 COVID-19 vaccine in adolescents: A randomized clinical trial. JAMA Netw. Open 2023, 6, e239135, doi:10.1001/jamanetworkopen.2023.9135.

6. Baden, L.R.; El Sahly, H.M.; Essink, B.; Kotloff, K.; Frey, S.; Novak, R.; Diemert, D.; Spector, S.A.; Rouphael, N.; Creech, C.B.; et al. Efficacy and safety of the mRNA-1273 SARS-CoV-2 vaccine. N. Engl. J. Med. 2021, 384, 403–416, doi:10.1056/NEJMoa2035389.

7. Dunkle, L.M.; Kotloff, K.L.; Gay, C.L.; Anez, G.; Adelglass, J.M.; Barrat Hernandez, A.Q.; Harper, W.L.; Duncanson, D.M.; McArthur, M.A.; Florescu, D.F.; et al. Efficacy and safety of NVX-CoV2373 in adults in the United States and Mexico. N. Engl. J. Med. 2022, 386, 531–543, doi:10.1056/NEJMoa2116185.

8. Heath, P.T.; Galiza, E.P.; Baxter, D.N.; Boffito, M.; Browne, D.; Burns, F.; Chadwick, D.R.; Clark, R.; Cosgrove, C.; Galloway, J.; et al. Safety and efficacy of NVX-CoV2373 Covid-19 vaccine. N. Engl. J. Med. 2021, 385, 1172–1183, doi:10.1056/NEJMoa2107659.

9. Polack, F.P.; Thomas, S.J.; Kitchin, N.; Absalon, J.; Gurtman, A.; Lockhart, S.; Perez, J.L.; Perez Marc, G.; Moreira, E.D.; Zerbini, C.; et al. Safety and efficacy of the BNT162b2 mRNA covid-19 vaccine. N. Engl. J. Med. 2020, 383, 2603–2615, doi:10.1056/NEJMoa2034577.

10. Alves, K.; Plested, J.S.; Galbiati, S.; Chau, G.; Cloney-Clark, S.; Zhu, M.; Kalkeri, R.; Patel, N.; Smith, K.; Marcheschi, A.; et al. Immunogenicity and safety of a fourth homologous dose of NVX-CoV2373. Vaccine 2023, 41, 4280–4286, doi:10.1016/j.vaccine.2023.05.051.

11. Sadoff, J.; Gray, G.; Vandebosch, A.; Cardenas, V.; Shukarev, G.; Grinsztejn, B.; Goepfert, P.A.; Truyers, C.; Fennema, H.; Spiessens, B.; et al. Safety and efficacy of single-dose Ad26.COV2.S vaccine against covid-19. N. Engl. J. Med. 2021, 384, 2187–2201, doi:10.1056/NEJMoa2101544.

12. San Francisco Ramos, A.; Liu Sanchez, C.; Bovill Rose, T.; Smith, D.; Thorn, N.; Galiza, E.; Miah, T.; Pearce, J.; Hultin, C.; Cosgrove, C.; et al. Comparing reactogenicity of COVID-19 vaccine boosters: A systematic review and meta-analysis. Expert Rev. Vaccines 2024, 23, 266–282, doi:10.1080/14760584.2024.2315089.

13. Centers for Disease Control and Prevention. Seasonal Influenza Vaccine Safety: A Summary for Clinicians. Available online: https://www.cdc.gov/flu/professionals/vaccination/vaccine_safety.htm (accessed on 4 June 2024).

14. Centers for Disease Control and Prevention. Shingles: About the Vaccine. Available online: https://www.cdc.gov/vaccines/vpd/shingles/hcp/shingrix/about-vaccine.html (accessed on 4 June 2024).

15. Gonen, T.; Barda, N.; Asraf, K.; Joseph, G.; Weiss-Ottolenghi, Y.; Doolman, R.; Kreiss, Y.; Lustig, Y.; Regev-Yochay, G. Immunogenicity and reactogenicity of coadministration of COVID-19 and influenza vaccines. JAMA Netw. Open 2023, 6, e2332813, doi:10.1001/jamanetworkopen.2023.32813.

16. Werner, F.; Zeschick, N.; Kuhlein, T.; Steininger, P.; Uberla, K.; Kaiser, I.; Sebastiao, M.; Hueber, S.; Warkentin, L. Patient-reported reactogenicity and safety of COVID-19 vaccinations vs. comparator vaccinations: a comparative observational cohort study. BMC Med. 2023, 21, 358, doi:10.1186/s12916-023-03064-6.

17. Rousculp, M.D.; Hollis, K.; Ziemiecki, R.; Odom, D.; Marchese, A.M.; Montazeri, M.; Odak, S.; Jackson, L.; Miller, A.; Toback, S. Burden and impact of reactogenicity among adults receiving COVID-19 vaccines in the United States and Canada: Results from a prospective observational study. Vaccines (Basel) 2024, 12, doi:10.3390/vaccines12010083.

18. Breeher, L.E.; Wolf, M.E.; Geyer, H.; Brinker, T.; Tommaso, C.; Kohlnhofer, S.; Hainy, C.; Swift, M. Work absence following COVID-19 vaccination in a cohort of healthcare personnel. J. Occup. Environ. Med. 2022, 64, 6–9, doi:10.1097/JOM.0000000000002376.

19. Nachtigall, I.; Bonsignore, M.; Hohenstein, S.; Bollmann, A.; Gunther, R.; Kodde, C.; Englisch, M.; Ahmad-Nejad, P.; Schroder, A.; Glenz, C.; et al. Effect of gender, age and vaccine on reactogenicity and incapacity to work after COVID-19 vaccination: A survey among health care workers. BMC Infect. Dis. 2022, 22, 291, doi:10.1186/s12879-022-07284-8.

20. Costa, K. Older Adults’ Intentions and Attitudes Toward the Updated Bivalent COVID-19 Booster 2023: Survey, United States, July 2023. Available online: https://www.healthcanal.com/health/the-bivalent-covid-19-booster-survey (accessed on 30 May 2024).

21. Rief, W. Fear of adverse effects and COVID-19 vaccine hesitancy: Recommendations of the treatment expectation expert group. JAMA Health Forum 2021, 2, e210804, doi:10.1001/jamahealthforum.2021.0804.

22. Freeman, E.E.; Strahan, A.G.; Smith, L.R.; Judd, A.D.; Samarakoon, U.; Chen, G.; King, A.J.; Blumenthal, K.G. The impact of COVID-19 vaccine reactions on secondary vaccine hesitancy. Ann Allergy Asthma Immunol 2024, doi:10.1016/j.anai.2024.01.009.

23. Tiozzo, G.; Louwsma, T.; Konings, S.R.A.; Vondeling, G.T.; Perez Gomez, J.; Postma, M.J.; Freriks, R.D. Evaluating the reactogenicity of COVID-19 vaccines from network-meta analyses. Expert Rev. Vaccines 2023, 22, 410–418, doi:10.1080/14760584.2023.2208216.

24. National Foundation for Infectious Diseases 2023 National Survey: Attitudes about Influenza, COVID-19, Respiratory Syncytial Virus, and Pneumococcal Disease. Available online: https://www.nfid.org/resource/2023-national-survey-attitudes-about-influenza-covid-19-respiratory-syncytial-vi rus-and-pneumococcal-disease/ (accessed on 12 March 2024).

25. Marchese, A.M.; Rousculp, M.; Macbeth, J.; Beyhaghi, H.; Seet, B.T.; Toback, S. The Novavax heterologous COVID booster demonstrates lower reactogenicity than mRNA: A targeted review. J. Infect. Dis. 2023, doi:10.1093/infdis/jiad519.

26. Munro, A.P.S.; Janani, L.; Cornelius, V.; Aley, P.K.; Babbage, G.; Baxter, D.; Bula, M.; Cathie, K.; Chatterjee, K.; Dodd, K.; et al. Safety and immunogenicity of seven COVID-19 vaccines as a third dose (booster) following two doses of ChAdOx1 nCov-19 or BNT162b2 in the UK (COV-BOOST): a blinded, multicentre, randomised, controlled, phase 2 trial. Lancet 2021, 398, 2258–2276, doi:10.1016/S0140-6736(21)02717-3.

27. Atmar, R.L.; Lyke, K.E.; Deming, M.E.; Jackson, L.A.; Branche, A.R.; El Sahly, H.M.; Rostad, C.A.; Martin, J.M.; Johnston, C.; Rupp, R.E.; et al. Homologous and heterologous Covid-19 booster vaccinations. N. Engl. J. Med. 2022, 386, 1046–1057, doi:10.1056/NEJMoa2116414.

28. Sutton, N.; San Francisco Ramos, A.; Beales, E.; Smith, D.; Ikram, S.; Galiza, E.; Hsia, Y.; Heath, P.T. Comparing reactogenicity of COVID-19 vaccines: A systematic review and meta-analysis. Expert Rev. Vaccines 2022, 21, 1301–1318, doi:10.1080/14760584.2022.2098719.

29. Salter, S.M.; Li, D.; Trentino, K.; Nissen, L.; Lee, K.; Orlemann, K.; Peters, I.; Murray, K.; Leeb, A.; Deng, L. Safety of four COVID-19 vaccines across primary doses 1, 2, 3 and booster: A prospective cohort study of Australian community pharmacy vaccinations. Vaccines (Basel) 2022, 10, doi:10.3390/vaccines10122017.

30. Stuart, A.S.V.; Shaw, R.H.; Liu, X.; Greenland, M.; Aley, P.K.; Andrews, N.J.; Cameron, J.C.; Charlton, S.; Clutterbuck, E.A.; Collins, A.M.; et al. Immunogenicity, safety, and reactogenicity of heterologous COVID-19 primary vaccination incorporating mRNA, viral-vector, and protein-adjuvant vaccines in the UK (Com-COV2): a single-blind, randomised, phase 2, non-inferiority trial. Lancet 2022, 399, 36–49, doi:10.1016/S0140-6736(21)02718-5.

31. Shaw, R.H.; Stuart, A.; Greenland, M.; Liu, X.; Nguyen Van-Tam, J.S.; Snape, M.D.; Com, C.O.V.S.G. Heterologous prime-boost COVID-19 vaccination: initial reactogenicity data. Lancet 2021, 397, 2043–2046, doi:10.1016/S0140-6736(21)01115-6.

32. European Medicines Agency. EMA Recommendation to Update the Antigenic Composition of Authorised COVID-19 Vaccines for 2024-2025. Available online: https://www.ema.europa.eu/en/documents/other/ema-recommendation-update-antigenic-composition-authorised-covid-19-vaccines-2024-2025_en.pdf (accessed on 26 May 2024).

33. World Health Organization. Statement on the Antigen Composition of COVID-19 Vaccines. Available online: https://www.who.int/news/item/26-04-2024-statement-on-the-antigen-composition-of-covid-19-vaccines (accessed on 26 May 2024).

34. Food and Drug Administration. Updated COVID-19 Vaccines for Use in the United States Beginning in Fall 2023. Available online: https://www.fda.gov/vaccines-blood-biologics/updated-covid-19-vaccines-use-united-states-beginning-fall-2023 (accessed on 26 May 2024).

35. Food and Drug Administration. Vaccines and Related Biological Products Advisory Committee June 5, 2024 Meeting Announcement. Available online: https://www.fda.gov/advisory-committees/advisory-committee-calendar/vaccines-and-related-biological-products-advisory-committee-june-5-2024-meeting-announcement (accessed on 26 May 2024).

